# The role of and key ingredients to community participation in health systems strengthening: a case study of the Matobo Village Health Sponsorship Model

**DOI:** 10.1101/2025.03.18.25324161

**Authors:** Tariro M. Chigwenah, Bhekimpilo Sithole, Chipo A. Nyamayaro, Faith R. Kandiye, Sikhululiwe Uzande, Thembelani Moyo, Takudzwa Mwanza, Cliff Zinyemba, Shamiso C. Madzivire, Mumo Sibanda, Mthabisi Msimanga, Andrew Muza, Norest Hama, Nomqhele Nyathi, Rudo M.S. Chingono

**Affiliations:** Independent Researcher; World Vision Zimbabwe, 59 Joseph Road, Mount Pleasant, Harare, Zimbabwe; Centre for Sexual Health HIV/AIDS Research, 4 Bath Road, Belgravia, Harare, Zimbabwe; Khomotayon Consulting Services, 80 Mutare Road, Harare, Zimbabwe; University of Zimbabwe, Department of Global, Public Health and Family Medicine, Harare, Zimbabwe; University of Zimbabwe, Department of Health Professions Education and Student Support Faculty of Medicine and Health Sciences; Biomedical Research and Training Institute, 10 Seagrave Road, Avondale, Harare, Zimbabwe; Ministry of Health and Child Care, Matebeleland Provisional Medical Directorate; United Nations Childrens Fund (UNICEF) Zimbabwe, Health Section Unit, 6 Fairbridge Avenue, Belgravia, Harare, Zimbabwe

**Keywords:** community participation, health systems strengthening

## Abstract

**Background:** Community participation is central to health systems strengthening and promoting healthcare provision in resource-limited settings. Several interventions have utilised community participation as a propellant to increase the access to and uptake of health interventions. However, while evidence supporting interventions using community participation exists, there is a gap in understanding the relative influence of various factors that enable or inhibit successful community participation. This study, which explored the enabling and inhibiting factors influencing community participation in the implementation of the Village Health Sponsorship Model (VHSM) in Matobo district addresses this gap. The VHSM is a primary healthcare intervention in Matobo, Zimbabwe, which by design and implementation relied heavily on community ownership and engagement.

**Methods:** A case study approach was employed to explore the enabling and inhibiting factors influencing community participation in the design and implementation of the VHSM. Data were collected in September 2022, in the form of key informant interviews (n=23), in-depth interviews (n=13) and participatory workshops (n=6). The data were audio-recorded and transcribed. The analysis process that followed included deductive coding and thematic analysis, both carried out manually.

**Results:** The socio-economic challenges in the Matobo community, along with limited availability and accessibility to healthcare services, and increased maternal, neonatal and child mortality were identified as key drivers fostering the community’s commitment to improving health outcomes in Matobo. Strong and effective community leadership, a sense of ownership, and a spirit of volunteerism were key to problem-solving. Locally sourced resources including manpower, water, and river sand contributed to the successful construction of three healthcare facilities, under the VHSM. The successful construction of the healthcare facilities in rural Zimbabwe provided evidence that community participation is an effective driving force towards achieving health systems strengthening in resource-limited settings.

**Conclusion:** Through community consultations, community involvement in resource sourcing, accountability of the community leadership, and public-private partnerships, three health facilities were constructed in the resource-limited setting of Matobo District. As we reflect on successful community participation driving the VHSM model and recommend this model for future interventions, we call for a shift towards health systems strengthening funding mechanisms that recognise communities as active agents of change.

## Background

Over the years, it has become apparent that there is an increase in the demand side of health systems, with the recent infectious disease outbreaks such as typhoid (1–3), COVID-19 (4,5), as well as the continuously high maternal and child mortality rates (6). This pressure on health systems, is a global concern that is further heightened in resource-limited settings. Partnerships across disciplines, sectors, and organisations are now essential for strengthening health systems. These collaborations enhance service delivery, financing, leadership, and more, while also supporting the health workforce (7,8). A core element of these partnerships is empowering individuals, families and communities to take charge of their own health (community participation), which has been framed as central to primary healthcare (9) and health systems strengthening (10). As both beneficiaries of, and key drivers to successful interventions, communities play an instrumental role in shaping and advancing health interventions that are contextually appropriate and relevant to their local needs (11).

Financial resources for health facility construction and health systems strengthening in low and middle-income countries (LMICs) are scarce and rarely allocated in alignment with community needs. As a result, LMICs have often relied on health systems financing and support from multinational donor agencies. Over the years, donor dependency has been reported to be high (12), and the sustainability of donor-driven interventions has increasingly become questionable. To curb this, there has been an increase in the promotion of community participation in primary health care programming. However, empirical evidence on the extent communities have participated, supported and driven such interventions is scant as the role and mechanisms of successful participation have not always been well documented (11,12). Of the studies that have documented successful community participation, key ingredients identified to participation include involvement throughout the process from the identification of the problem, the identification and development of the interventions, the involvement in the implementation, resource allocation and monitoring and evaluation of the intervention (13,14).

Considering the limited funding available for health systems establishment and health systems strengthening, achieving sustainable health systems strengthening in resource limited settings like Zimbabwe would require a shift from donor driven funding to recognising local stakeholders and communities as equal players in the improvement of community level health outcomes, combined with increased domestic financing. The objective of this study was to understand the nature, extent, and quality of community participation in the Village Health Sponsorship Model (VHSM) implemented as a health systems strengthening intervention in Matobo District in Zimbabwe.

The VHSM model was established as an afterthought of how best to utilise remaining funds of World Vision Zimbabwe’s (WVZ), Matobo District Area Program (under its Development Program Approach). World Vision Zimbabwe had been operating in Matobo’s for 11 years and during this operational period, WVZ realised that the village was contributing significantly to the health facility construction projects. Following this realisation, a request was sent to the donor that the remaining funds (for the Area Programme) be diverted to supporting and serving the communities’ identified as in need of healthcare facilities. Acknowledging the communities as agents of change equipped with key resources (such as local capacity and leadership), WVZ, aligned to its transformational development principles (15,16), developed the Village Health Sponsorship Model. As part of this model, they supported the construction of three healthcare facilities through the strengthening of local capacity and leadership, partnerships, and the provision of additional financial support.

Recognising the importance of assessing impact in a way that expands the evidence base on effective collaborative efforts between various sectors, this study seeks to evaluate the role of community ownership and participation in the construction of the three health facilities using the World Vision Zimbabwe (WVZ) Village Health Sponsorship Model. This research is pertinent as it provides an in-depth exploration of the role of community participation, and the viability of the VHSM as a model for increasing healthcare facilities in order to strengthen health systems in resource-limited settings.

## Methods

### Study setting

This case study was conducted in September 2022I n three rural primary healthcare facilities in Matobo District namely, Ndabankulu, Fumugwe and Silozwi. Matobo District was purposefully chosen as this is where the VHSM model was established. The district is predominantly a rural area, with limited accessibility to healthcare facilities, educational facilities, transport and other basic amenities. Matobo District had an enumerated population of 95 696 people in 2022 (17). The district’s referral system consists of two tertiary healthcare facilities (one government hospital and one mission-funded hospital), both situated over 20 kilometres from the chosen primary healthcare facilities.. The vast distance between communities and the referral hospitals has motivated communities to partner with other stakeholders to construct community level health facilities. The three communities are in a drought-stricken area. Economically, Ndabankulu is a mining community, and the other two are sustained by diaspora remittances and local businesses.

### Study design and population

The study is a case study which employs qualitative research methods to allow for an in-depth understanding of the role of community participation in the successful construction of healthcare facilities through the VHSM. The study targeted key stakeholders of the VHSM in the particular communities. Stratified purposive sampling was employed to select respondents from all three facilities who were believed to be better positioned to give insight into the background, implementation and learning points of the VHSM. 25 key informants (government line ministries, community leaders, i.e., chiefs, village heads, health committees leads, and village health workers) were targeted. 15 community beneficiaries were targeted as active agents and beneficiaries of the VHSM.

### Theoretical Framework

This study drew upon Chaskin (2001)’s theory of community capacity to assess the means and level of community ownership and engagement in the VHSM. Key elements of this theory are: i) a sense of community (based on mutual values, concerns and benefits); ii) commitment among community members (based on concern for the general wellbeing of the community); iii) mechanisms of problem solving (having identified a common problem/goal and agreed upon how to take it); and iv) access to resources (based on what the community can bring to the table to establish the change they desire) (18). This study theorises that the VHSM was successful as it had these fundamental elements needed to foster community ownership and full community engagement.

### Data collection and analysis

Data collection was conducted in September 2022 by five research assistants and a health economist, under the supervision of a Social Scientist (first author). Two key qualitative research methods were used, namely in-depth interviews and participatory workshops.

Topic guides were developed for the in-depth interviews (n=40) and participatory workshops (n=6). These included questions on the key factors leading to the conceptualisation of the facilities, the intricate details of the process, perceived enablers and barriers during the construction phase, and the perceived impact of the rural health clinics, and assessing the role of community participation in leadership, resource mobilisation, and sustainability of the model. The interviews were conducted in the local language of preference, with majority of the interviews being in Ndebele. Interviews ranged from 20 - 60 minutes. Two participatory workshops were conducted in each community, one with community beneficiaries and another with key informants. Workshops ranged from 90 – 120 minutes. To better understand the role of the community, the in-depth interviews and participatory workshops tried to elicit the societal perspective on the various forms of contributions various partners of the VHSM made to the construction of the facility.

The above data was audio-recorded and transcribed verbatim and then translated into English. The transcripts were manually coded, and thematic analysis was employed to analyse the conceptualisation and construction of the healthcare facilities, and the role of community participation in the successful implementation of the VHSM. Codes and themes developed were informed by Chaskin (2001) theory of community capacity. FK and CN independently coded a sample of eight transcripts simultaneously. They discussed the outcome of their independent coding with RMSC and deductively identified the themes and codes that were related to Chaskin’s theory as well as other codes. The remaining transcripts were then coded based on the developed framework, which was refined as coding continued.

### Ethics

The study obtained regulatory approval from the Medical Research Council of Zimbabwe (MRCZ), reference number MRCZ/A/2960. Participants were informed of the study, its procedures, minimal risks and benefits. Written consent was sought from the study participants.

## Results

### Summary of results

A total of 23 key informants, and 13 community members took part in the in-depth interviews. Six workshops and three health facility surveys were conducted as part of this study. Key themes and codes include

i. contextual factors creating the need for improved health systems (inaccessible healthcare facilities, high maternal and child mortality rates, transport challenges to access health facilities), restricted access to essential medicines,
ii. enablers of active community participation (good and effective community leadership, community buy-in and a sense of ownership, capacity to contribute resources, capacity building and private public partnerships),
iii. and barriers to community participation (lack of trust, volunteer attrition, competing demands).

Figure 1 below highlights some of the key elements towards active community participation that led to the successful implementation of the health facilities using the VHSM.

**Figure 1:**
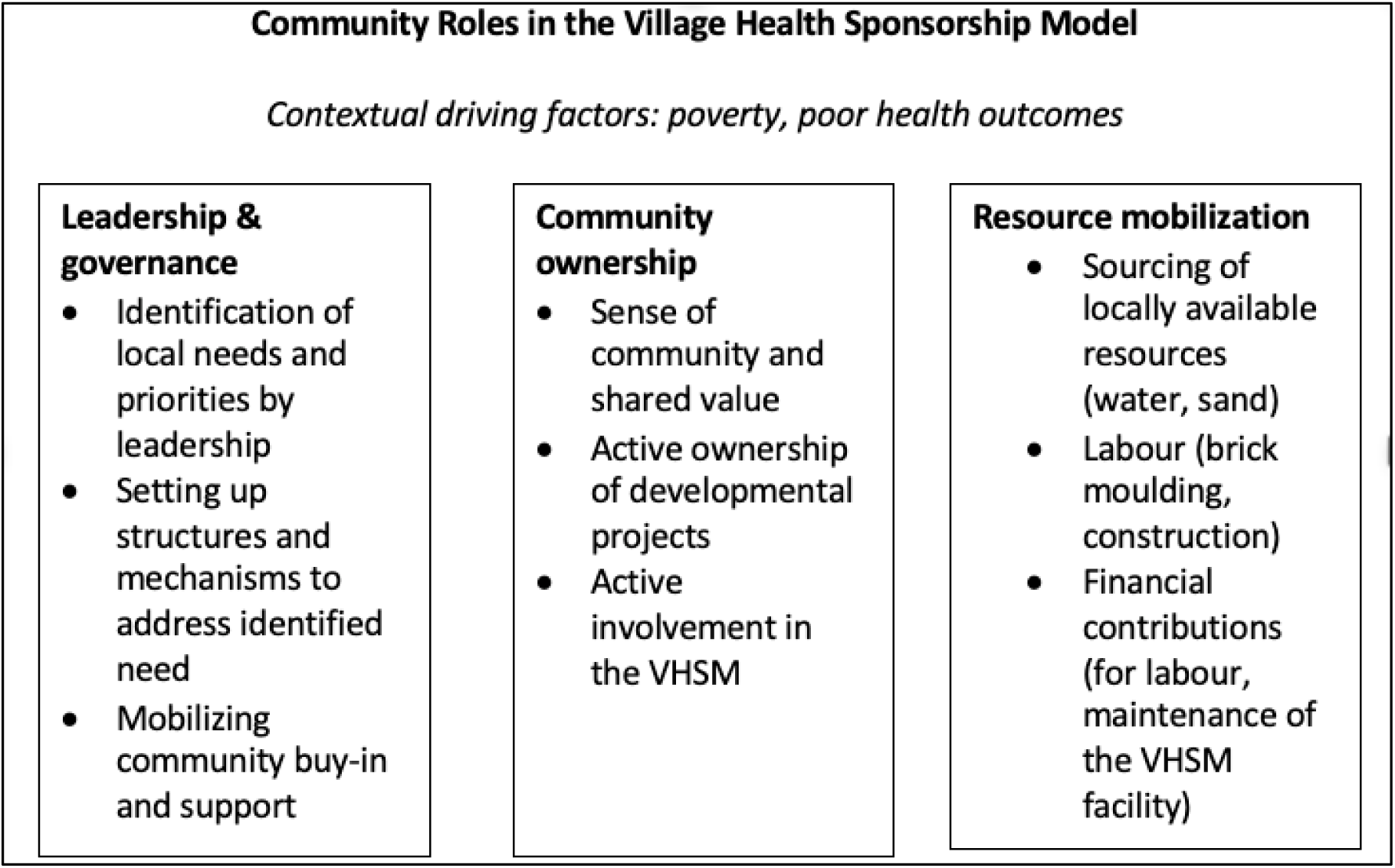
Active ingredients to the community participation in implementing the Village Health Sponsorship Model.

### The Matobo Village Health Sponsorship Model – Conceptual Framework

Figure 2 shows the Matobo VHSM conceptual framework established from the results from the data collected during the study. Founded on the principles of transformational development, the model acknowledged the role of the community in identifying problems within their context, that resulted in undesired health outcomes. To address these undesired health outcomes, the three communities prioritised constructing health facilities to reduce the distance and increase access and uptake of healthcare services. The belief was that this would lead to improved health outcomes in the long run. To facilitate the construction of the healthcare facilities, partnerships were established between community members, local businesses, local children in the diaspora, development partners, and government ministries. Of particular focus to this conceptual framework is community participation, and its active ingredients (labelled 1, 2, and 3 in Figure 2) that had been previously identified in Figure 1.

**Figure 2:**
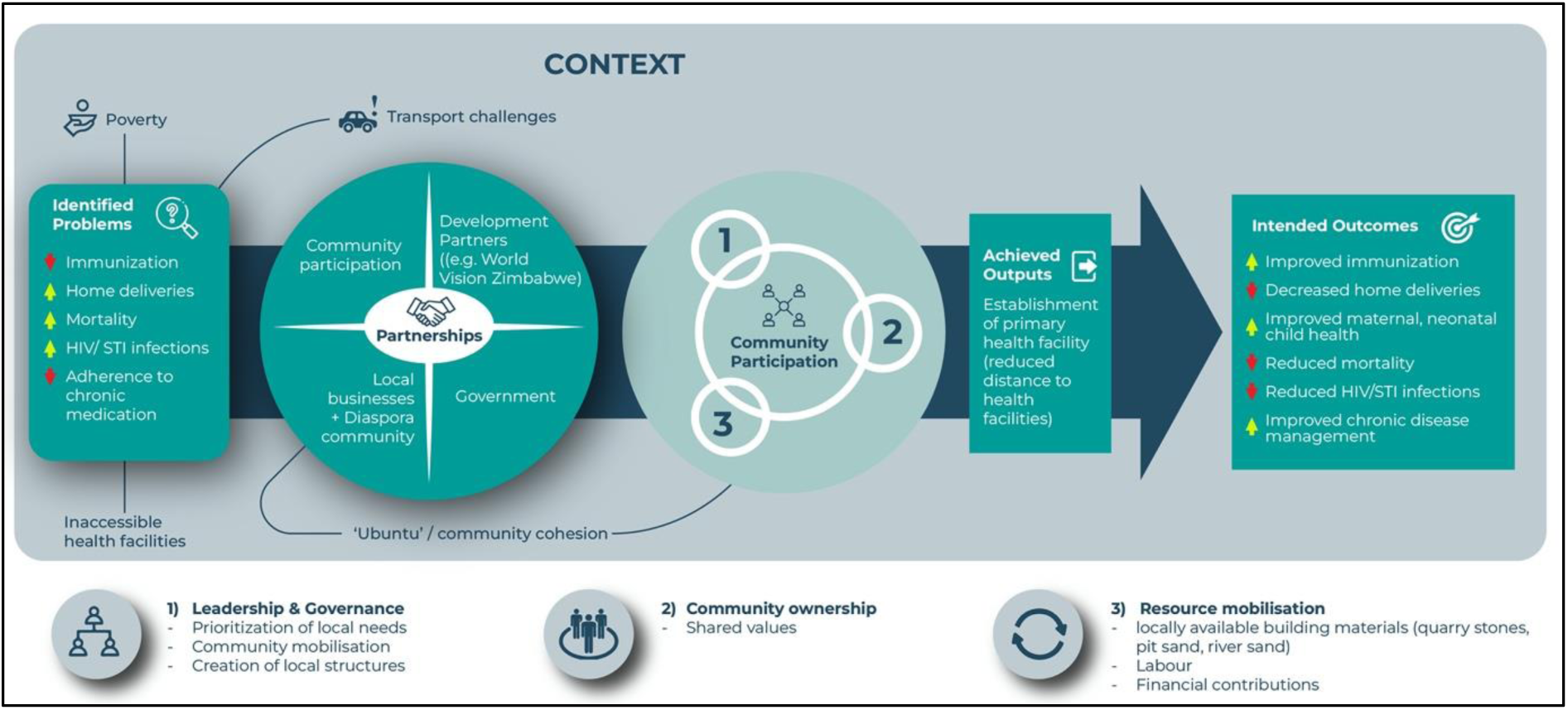
Conceptual Framework of the Matobo District Village Health Sponsorship Model. Figure 2 shows the VHSM conceptual framework. The model describes the establishment of partnerships in a context where poverty is high, distance to health facilities is long, transport is scarce and expensive, making access to basic services like healthcare inaccessible. Particular health challenges were identified as emanating from contextual challenges, and in an attempt to address these, partnerships were formed with several key players. The framework zones in on the role of community partnerships, three key ingredients (leadership and governance, community ownership, resource mobilization) that contributed to the desired outputs (three clinics), and intended (improved health) outcomes.

### Contextual factors creating the need for improved health systems

Findings from the study showed that prior to the VHSM, there were contextual factors that affected access and uptake of healthcare services. There were limited and widely distributed healthcare facilities, making healthcare services inaccessible, consequently resulting in a negative impact on health outcomes including poor maternal health, increased home deliveries, lack of child immunisation and increased mortality.

#### Inadequate healthcare financing

Lack of adequate health financing in Zimbabwe was revealed as a central problem that affected Matobo communities, like most rural communities in the country. There are limited local government resource allocations that trickle down to enable district and local governance to build health facilities that are accessible and fully equipped.

> “Our health needs as a country have surpassed our national budget for health for our growing population. Our health systems are under-resourced, we do not have adequate infrastructure, medicine, and of late our personnel is being depleted with everyone leaving for the diaspora…here in Matobo we only have 1 public hospital” (Key informant_22_Male)

#### Inaccessible healthcare services and poor health outcomes

Access to healthcare services was restricted and this was largely due to healthcare facilities being distant, transport being scarce and costly for people to travel.

> “People used to board transport to the hospital that is far which is about 25km, when you dropped off you would walk another 5km but those who had money would board again another mode of transport to reach the hospital.” (Community member_03, Female, Clinic 01)

> “If we look at the distance that is travelled going to district hospital, it is too far and expensive one would need R50 (approximately US$3) to and R50 from… It was very expensive because one needed food and also if the medication was not available in the hospital you would be forced to buy [from a private pharmacy]. At the end of the day people were being forced to sell their livestock to access healthcare services.” (Key informant_08, Male, Clinic 02,)

> “Then another challenge is transport. If you want to accompany or visit your spouse in hospital, you will end up not doing so… The challenge is money for travelling as we board twice. One also worries about where you will spend the day or where you will sleep as it is far, and you will want to avoid commuting daily.” (Key informant_11, Male, Clinic 02,)

This resulted in communities seeking health services primarily, only, in the case of emergencies. This more often than not resulted in adverse health outcomes including increased home deliveries, and disease outbreaks.

> “Three years back I could not go to the clinic because it was very far, I couldn’t afford the transport, so I would heal from home purchasing painkiller tablets from the shop and sometimes I would be given tablets from neighbours and friends not knowing their purpose just taking the pills.” (Community member_01, Female, Clinic 01)

> “Children who were supposed to go for immunisation and growth monitoring never got immunised on time …When there was an outbreak like measles people would get sick and some would die because there was no clinic available close. The greatest challenge we also faced was that we lived near a river that had no bridge for people to cross over so we would go with nursing mothers to a nearby Secondary School so that they could access the mobile baby clinic.” (Village health worker 04, Female, Clinic 03)

### Enablers of active community participation

#### Good leadership and governance

Community leadership was identified as a key contributor to the success of the VHSM. The qualitative data showed that the community leadership made efforts in identifying the community health needs and mobilised villagers and local business partners to pave a way forward in improving access to health services. Key attributes that made leadership viable to implement the VHSM in these three communities include; having the community interests and needs at heart, accountability and trustworthiness.

> “If you are a leader you should have your people at heart, do things that are beneficial to everyone, you should engage with the people don’t exercise your power too much. You can be a chief but also learn how to live with people despite your post. This will help you win their trust and following. If they don’t trust you … coordinate and use someone else whom the people respect and then work together, let him talk to the people.” (Key informant_11, Male, Clinic 02)

> “Delegate duties in order to have equitable accountability, leave the duties to other people so they can monitor what is going then you can correct.” (Key informant_12, Female, Clinic 02)

#### Community buy-in and ownership

Community members’ buy-in to the vision and proposed development plans played an intricate role in the success of the VHSM. It is perceived that community members’ awareness of the problems of distance to the health facility and the impact of failed access to health services, such as home deliveries, maternal deaths, and disease outbreaks, made it easier for the community members to take heed of the call to construct a local health facility.

> “The long distance, the death of children due to giving birth at home sometimes they were special cases that needed attention but you would get transport late and the time you reach the clinic the child is dead…all of these made it easier for the community to want to ensure change.” (Community member_03, Female, Clinic 01)

> “One of the major reasons why we built the clinic was because we were worried about pregnant women who had to travel long distances to Bhazha clinic, and some could not afford to board a bus.” (Key informant_13, Male, Clinic 03)

#### Capacity to contribute resources

Another key enabler to the success of the VHSM, was believed to be the capacity of the community members to contribute towards the required resources for the construction of the health facility. Community leadership and the health committees calculated the resources needed for the construction and these were distributed to each village and household. Resources contributed by the community were labour (for moulding bricks and constructing the facility), water, river sand, pit sand, and quarry stones. They also made financial contributions.

> “Yes, the community contributed all of the bricks (250 bricks per household), quarry, river sand, pit sand and even the mixing of cement they helped in the manual labour as well.” (Key informant_08, Male, Clinic 02)

> “We were responsible for brick moulding, looking for quarry, river sand and containers. We brought the quarry stones and river sand walking 2km, and also the bricks were far about 500m so we carried them with bare hands to bring them close.” (Community member_01, Female, Clinic 01)

> “When the time came for us to start the building project, as a burial society we asked if it was possible for us to take the money that had been set aside, which we would replace when the community contributed. When the community made contributions and even the ones in South Africa (diaspora) made money contributions, we managed to replace the money.” (Key informant 19, Male, Clinic 03)

#### Capacity building and public-private partnerships

The results show that public-private partnerships played a fundamental role towards the successful implementation of the VHSM, as participants expressed the several funding constraints, they had that stalled the progress of their infrastructure development. It was cited that it was through public-private partnerships between the community and its local leadership, the rural district council, development partners (such as World Vision Zimbabwe), local business partners and family members living in the diaspora that the three communities managed to build their health facilities.

> “Community members were very supportive they brought other building materials such as sand and bricks… There was a miner who wanted to help us because they had heard that people wanted to build a clinic. He donated cement, fence and poles for fencing…. those who did plumbing were local people like us. The things were not enough to finish the work but that is when World Vision came and helped. The people who did the ceiling were from Plumtree and were brought here by the World Vision.” (Key informant_08, Male, Clinic 02)

> “The children (locals in the diaspora) had come over here for the holiday. They coordinated with the councilor who asked for assistance in their project, and they accepted…they really gave a hand in the project. What I noticed myself is that they paid for the bricks for us.” (Key informant_10, Male, Clinic 02)

> “As a member of the local leadership I noticed the construction material was not enough, so we mobilized transport. The National Park was asked for a tractor to help us carry sand, we also asked Ebenezer they are close by here, so they also helped in carrying the sand.” (Key informant_01, Male, Clinic 01)

### Barriers to community participation

The study findings did highlight that despite the successful implementation of the VHSM, there were challenges in implementing this model. Barriers encountered during the implementation of the VHSM include feelings of distrust in and resistance to community leadership, financial constraints and volunteerism attrition.

#### Lack of interest, distrust, and resistance to local leadership

Findings show that there was resistance or lack of participation by other community members who did not contribute to the resources as per expectation, whilst others committed on paper but did not honour their commitment of labour or resources.

> “They were refusing, some said we have never seen a clinic being built from mud bricks, some would say I won’t waste my time going for the construction and others said it was all politics so we don’t do that, till the clinic got finished some didn’t even come not even a single day…They are the ones in the forefront now (laughs).” (Community member_02, Female, Clinic 01)

> “When we started many people were in support of the idea but practically, they did not show up in numbers because when it comes to manual labour and things that need money people tend to not cooperate.” (Key informant 06, Clinic 03, Male)

Some of the reasons cited for lack of community participation included the distrust in local leadership. If a person did not “trust the leader, or they had a personal grudge, or they were from a political party not aligned to theirs” that had a negative impact on the participation of the community member.

> “What I can say is that what affects other regions is politics. People should focus more on development and not politics. Those in power, like chiefs, should also coordinate others they shouldn’t be power greedy” (Key informant, Clinic 02, Male)

#### Financial constraints

Financial constraints posed a major challenge that hindered community participation especially in the form of financial support. In the three different communities, households were assigned certain amounts at certain stages of the construction, but not every household was able to contribute as agreed upon.

> “People agreed to contribute as per household and each household was supposed to contribute 20 US dollars or the equivalent of that in bond Zimbabwean currency. Unfortunately, it was not a success because people had financial challenges.” (Key informant_08_Male, Clinic 02)

This posed as a major delay in construction as the lack of financial contribution meant builders who were meant to be paid in cash or given a token of appreciation did not end up getting this resulting in volunteerism attrition.

> “We had a challenge contributing money to give the builders. The council had promised that they will give them wages, but they failed to raise them and at the end of the day they stopped” (Key informant_12, Male, Clinic 02)

> “Everyone who is a builder got their names noted down…but when it came to action only a few came (both laugh) because of influences from others and if for instance, a wife complained that they are suffering from hunger at home whilst he spent time at the clinic working with no wage those men ended up retreating leaving only a few.” (Community member_05, Female, Clinic 01)

### Positive outcomes from the VHSM

The VHSM was perceived to be a successful model in health systems strengthening, as the three communities in which it has been implemented were able to successfully build primary healthcare facilities. The results of this study highlighted positive outcomes resulting from community participation in the construction of the healthcare facilities.

#### Improved health outcomes

Improved health outcomes were perceived to be a major outcome identified across the three communities. It is perceived that there has been a shift of seeking health services in dire situations or emergencies, to proactive health seeking behaviour. People with chronic conditions were reported to have increased access to their chronic medication and this was believed to have improved adherence to treatment and disease management. Child immunisation was believed to have increased and consequently it was reported that there was perceived reduction in disease outbreak.

> “Now the greatest advantage we have is that we have reduced number of illnesses and even complications because people get help faster. Now they are reduced home deliveries because back then some women will delay going to the hospital and end up giving birth along the way to hospital.” (Key informant_08, Male, Clinic 02)

> “As of now there are no defaulters anymore because a person would fail to get someone to send and get tablets for them at the clinic… but now it’s easier because the clinic is now near.” (Community member, Female, Clinic 01)

> “The number of deaths in children has greatly reduced, because children would die because they were not immunised.” (Key informant_21, Female, Clinic 03)

#### Sustainability of community-led initiatives

The data shows that through the successful construction of the health facilities using the VHSM, community unity strengthened in all three communities. The sense of achievement triggered the desire to address more community needs.

> “This initiative has made people work together. It has motivated people and as of now they want to construct a cottage for the nurses because they have realised working together works. Thanks to that they have learned that working together is an easier way they can fend for themselves.” (Key informant, Male, Clinic 02)

In some of the communities there was a continuation of the development initiatives as community members went on to commit to addressing other gaps they identified. In one community they had gone on and started constructing a waiting mothers’ shelter, in another they had come up with a community garden to help feed the healthcare workers and those clinic patients identified to need nutritional support.

> “Despite the resistance at first, we have all witnessed the benefit of uniting in our community development. This is our community, we need to develop it. Even those who were backbenchers are asking how shall we contribute (laughs)…Now we are working towards building the waiting mothers shelter. As you can see the bricks are now trickling in.” (Key informant_05, Male, Clinic 01)

> “We have established a community garden, that will help to feed the mothers in the waiting mothers’ shelter as well as help feed our nurses. People take turns to come and water. Now there are talks of raising resources to build another house for the nurses.” (Community member_07, Male, Clinic 02)

### Recommendations for future development plans

Despite its success, the challenges faced whilst implementing the VHSM, influenced the respondents’ perception on the importance for *Ubuntu -* emphasizing the interdependence of people and the need for communities working together towards the development and advancement of their health systems. Traditional leaders were also called upon to play their roles as community focal persons, ensuring that community members participate fully in VHSM initiatives in order to make implementation successful.

> “As a community you need to sit down together [ubuntu] and raise all the concerns you have because at the end of the day it’s you who suffer by walking long distances for a service that you can build when you unite and gather resources.” (Key informant_08, Male, Clinic 02)

> “Through our leadership, we have been able to rally the community. Those that have tried to rally together people but have failed it is because the leadership, which forms the foundation, is not strong… I would recommend those wanting to work towards a community project should make sure their leaders are in good books with their community… they should have the community at heart… that way people will listen to them.” (Community member_13, Male, Clinic 03)

> Capacity building of local communities was identified as a key recommendation for successful community participation and ownership. A major gap in community participation that was identified was record keeping. Community members need to be shown how to document their meetings, how to document the resources brought in and utilised for such initiatives.

> “On record keeping there is a need for people to be educated in all committees so they know the importance of record keeping, because if we look deep into it when I arrived some materials had arrived already but to trace it back to who contributed and gave a hand for instance in the diaspora it cannot be traced. So on transparency there is still a challenge, there is need for trainings.” (Key informant_01, Male, Clinic 01)

The data from the interviews shows that some community members and leadership believe that the concept of the VHSM can be applied towards the attainment of any development plans the community sets out to achieve, for example, the construction of schools, nutrition gardens, and community boreholes.

## Discussion

The objective of this study was to understand the nature, extent, and quality of community participation in the Village Health Sponsorship Model implemented as a health systems strengthening intervention in Matobo District in Zimbabwe. This study showed that for the implementation of the VHSM in Matobo to be successful, community ownership and participation was a core feature. This study has provided the key ingredients of community engagement in the VHSM, with the ultimate aim of informing future health strengthening initiatives. A major health outcome of the VHSM is the expansion of health service availability in the communities. The improved geographical accessibility to a health facility was perceived to have resulted in improved access to maternal neonatal child health services, improved access to chronic disease testing and treatment, increased access to family health services and consequently reduced morbidity and mortality.

To fully conceptualise these dynamics of community participation, this study situated the analytical approaches to successful community participation within Chaskin’s (2001) theory of community capacity (18). Chaskin’s theory of community capacity recognises the sense of community and unity as an “active ingredient” to successful community engagement (18). Aligned to this, there are several key ingredients identified (see **Error! Reference source not found.**) for the successful buy-in and engagement of community members resulting in the construction of the three health facilities in Matobo District. These include good leadership and governance, the sense of community and community members commitment to addressing their developmental needs, the capacity to provide resources, public-private partnership to cite but a few. A review conducted by Singh et. al., (2017) highlights how several other studies have shown evidence of how community participation has a positive impact on health; as health outcomes can be improved when communities identify with and claim ownership of the development initiatives (19). In our study, we found that through good local leadership and governance, the three communities had a strong sense of community. They were able to unite in identifying their local needs and work together to the point of the successful construction of local healthcare facilities. This highlights how development initiatives, led or conducted under the guidance and support of local leadership, are more likely to be successful than those imposed upon communities. Similarly, a study by Baltzell, et. al., (2019) identified community engagement, particularly community leaders involvement (as key influencers), in the design and execution of development initiative to be critical in the drive towards malaria elimination in several contexts (20).

Community engagement is most successful when engagement is iterative and responsive to changing community needs, perceptions, and opinions. Aligned to Chaskin’s theory, this study showed, in Figure 2, that where community needs are identified, addressed, and their general well-being promoted, there is commitment in community participation. Establishing the problem or need, and getting community buy-in on possible mechanisms of problem solving was another key component that Chaskin postulated enabled community capacity to engage (18). Similarly, other studies have previously identified community participation as a key component in interventions where population health improved (21).

The importance of public-private partnership in the successful construction of healthcare facilities in Matobo District was highlighted as a key attribute to enhancing community participation in this study. The construction of the healthcare facilities was fast-tracked by the embedded presence of organisations, business partners, and family members in the diaspora who were willing to support the community’s vision and efforts. Additional key components of the success experienced are those of the bidirectional communication that occurred in the partnerships, and the presence of trust built over the longstanding engagements between the community and its development partners. This study illustrated a mechanism that is feasible for communities, health sector actors and other sector actors to gather and respond to different local health and development needs. Aligned to the findings, a study by Maat et. al., (2021) highlighted four case studies portraying the important role of public-private partnerships in co-producing locally relevant response to health issues (22). In Madagascar, the combined community and NGO efforts to the Hurricane emergency health responses allowed for continuity of basic healthcare support, minimised disease outbreaks, and improved food security. In Uganda, university research and training of locals helped support the construction of community-developed sanitary facilities (22).

This study has shown that although the communities successfully built the healthcare facilities, they did encounter problems in community participation highlighted through some community members failing to contribute as agreed upon. This resonates with earlier studies that have shown that individualism, apathetic behaviours and weak communal ties can result in a low sense of community which can ultimately result in failure or the setting back of the successful implementation of development initiatives (23).

To prevent setbacks and promote sustainability of local participation, the VHSM conceptual framework highlights fundamental ingredients for this. Community participation, mobilisation of resources by the community, authentic partnerships, community identified mutual benefit can encourage the sustainability of strengthening local health systems. A study in Zambia showed that involvement of community members can increase the uptake of voluntary medical male circumcision programmes. Additionally, leveraging in already existing community structures may reduce the general costs of implementing the programme (24). Another study conducted in rural Nepal revealed that the successful implementation of community-led safe water infrastructure fostered a sense of ownership that encouraged the successful management of a shared resource (13). Community participation, that results in successful outputs, can lead to the feeling of psychological ownership and can enhance stewardship behaviour that promotes the sustainability of strengthening local health systems (13).

### Strengths and limitations

Promotion of community participation has been accelerated in health system strengthening activities, but scarce evidence exists about what makes community participation a successful driving force. A major strength of this study is that it provides insights into the various roles the community played and the key ingredients to the success of community participation in the construction of healthcare facilities. In the VHSM, communities have been shown to play a vital role in planning, priority setting, model implementation and expansion.

A major strength of this study is the in-depth understanding it provides, and conceptualisation of the Village Health Sponsorship Model (Figure 2), which can be applied to varying degrees to different contexts and can be adapted for addressing different Sustainable Development Goals (such as construction of schools under SDG# 4). This conceptual framework is not exhaustive, but it makes valuable contribution to the health systems strengthening debate, particularly highlighting the importance of communities, their agency and resources in achieving universal health coverage.

Despite the strengths and important findings made in the current study, it is worth mentioning its potential limitations. As a qualitative study, our research relied on the self-reported views of participants for discussions and conclusions. While we recognise that appropriate stakeholders and community beneficiaries were involved in the interviews and participatory workshops, their personal biases could have influenced the views expressed. The participatory nature of the workshops in the study, however, ensured that consensus was reached on the major challenges in the implementation of the VHSM.

Another gap is that data was collected retrospectively, after the implementation of the VHSM and the successful construction of the healthcare facilities. During data collection it was evident that there was a lack of documentation of community contribution towards the implementation of the VHSM in the construction of the healthcare facilities. Data collected was therefore restricted to people’s recollection of events and where available a few documented records by the health committee secretary. Addressing such gaps in raising evidence of community participation in health systems strengthening will require creating tools or standardised reporting systems that will facilitate the evaluation and documentation of community services. These tools can be developed for community training as part of capacity building provided through the public-private partnerships.

## Conclusion and recommendations

Evidence from this case study suggests that community participation is a feasible strategy for health systems strengthening in resource-limited settings. While there are several papers that explore the role of community participation in health systems strengthening in resource-limited settings, this study offers several conclusions to expand on that exploration, as well as insight into the “active ingredients” of successful community engagement. Key ingredients to active community engagement leading to the success of the VHSM include but are not limited to, the early involvement and buy-in of community leadership and community members, having common interests and goals, the capacity to source resources by community members and, having functional public-private partnerships.

Future programmes need to support communities in the processes and accountability mechanisms in decision-making, and the implementation and documentation of development initiatives using approaches such as the VHSM. There is need for further research that uses a wider pool of data to identify the support and investments necessary to empower favourable characteristics for community participation, and promote more robust community-based health systems. In conclusion, this study has shown that construction of healthcare facilities in resource-limited settings, is achievable and health systems strengthening can be spearheaded through community participation in partnership with local and international development partners.

## Supporting information

Supplemental table of key themes

## Data Availability

All data produced in the present study are available upon reasonable request to the authors

