## Supplemental table of key themes for "The role of and key ingredients to community participation in health systems strengthening: a case study of the Matobo Village Health Sponsorship Model"

| Theme | Sub-theme | Quotation |
|---|---|---|
| Contextual factors creating need for improved health systems | Inadequate health financing | |
| | Inaccessible healthcare services and poor health outcomes | *“People used to board transport to the hospital that is far which is about 25km, when you dropped off you would walk another 5km but those who had money would board again another mode of transport to reach the hospital.” (Community member, Female, Clinic 01)* |
| | | *“You see that back in the day one would travel to Natisa whilst you are sick, so by the time you reach the clinic your sickness would have worsened, by the time you reach there you can’t even walk (laughs). So it is really good that the clinic is now close” (Key informant, Male, Clinic 02)* |
| | | *“Three years back I could not go to the clinic because it was very far, I couldn’t afford the transport, so I would heal from home purchasing pain killer tablets from the shop and sometimes I would be given tablets from neighbours and friends not knowing their purpose just taking the pills.” (Community member, Female, Clinic 01)* |

| | | |
|---|---|---|
| | | *“When there was an outbreak like measles people would get sick and some would die because there was no clinic available close. The greatest challenge we also faced was that we lived near a river that had no bridge for people to cross over so we would go with nursing mothers to a nearby secondary so that they could access the mobile baby clinic.” (Village health worker 04, Female, Clinic 03)* |
| | | *“children who were supposed to go for immunization and growth monitoring never got immunized on time. Then all that brought complications in that a child would fall sick due to measles and when their cards were checked one would discover that the child was never immunized.” (Village health worker 04, Female, Clinic 03)* |
| | Transport and logistics costs and challenges | *“If we look at the distance that is travelled going to Maphisa, it is too far and expensive one would need R50 to and R50 from… It was very expensive because one needed food and also if the medication was not available in the hospital you will be forced to buy. At the end of the day people were being forced to sell their livestock.” (Key informant_08, Male, Clinic 02)* |

| | | |
|---|---|---|
| | | *"Then another challenge is transport. If you want to accompany or visit your spouse in hospital you will end up not doing so… The challenge is money for travelling as we board twice. One also worries about where you will spend the day or where you will sleep as it is far, and you will want to avoid commuting daily." (Key informant, Male, Clinic 02)* |
| Enablers of active community participation | Good leadership and governance | *"If you are a leader you should have your people at heart, do things that are beneficial to everyone, you should engage with the people don't exercise your power too much. You can be a chief but also learn how to live with people despite your post. This will help you win their trust and following. If they don't trust you … coordinate and use someone else whom the people respect and then work together, let him talk to the people." (Key informant, Male, Clinic 02)* |
| | | *"Delegate duties in order to have equitable accountability, leave the duties to other people so can monitor what is going then you can correct." (Key informant, Female, Clinic 02)* |
| | Community buy in and ownership | *"The long distance, the death of children due to giving birth at home sometimes they were special cases that needed attention but you would get transport late and the time you reach the clinic* |

| | | |
|---|---|---|
| | | *the child is dead…all of these made it easier for the community to want to ensure change ” (Community member, Female, Clinic 01)* |
| | | *“One of the major reasons why we built the clinic was because we were worried about pregnant women who had to travel long distances to Bhazha clinic, and some could not afford to board a bus.” (Key informant 06, Male, Clinic 03)* |
| | Capacity to contribute resources | *“Yes, the community contributed all bricks (250 bricks per household), quarry, river sand, pit sand and even the mixing of cement they helped in the manual labour as well.” (Key informant_08, Male, Clinic 02)* |
| | | *“We were responsible for brick moulding, looking for quarry, river sand and containers. We brought the quarry stones and river sand walking 2km, and also the bricks were far about 500m so we carried them with bare hands to bring them close” (Community member, Female, Clinic 01)* |
| | | *“We used to carry water from the dam and women would use their heads to carry water. Things got better when a tap was drilled.” (Key informant_08, Male, Clinic 02)* |

| | | |
|---|---|---|
| | | *"When the time came for us to start the building project, as a burial society we asked if it was possible for us to take the money that had been set aside, which we would replace when the community contributed. When the community made contributions and even the ones in South Africa made money contributions, we managed to replace the money." (Key informant 06, Male, Clinic 03)* |
| | Capacity building and private public partnerships | *"Community members were very supportive they brought other building materials such as sand and bricks… There was a miner who wanted to help us because they had heard that people wanted to build a clinic. He donated cement, fence and poles for fencing…. those who did plumbing were local people like us. The things were not enough to finish the work but that is when World Vision came and helped. The people who did the ceiling were from plumtree and were brought here by the World Vision Organisation." (Key informant_08, Male, Clinic 02)* |
| | | *"The children (locals in the diaspora) had come over here for the holiday. They coordinated with the councilor who asked for assistance in their project, and they accepted…they really gave* |

| | | |
|---|---|---|
| | | *a hand in the project. What I noticed myself is that they paid for the bricks for us." (Key informant, Male, Clinic 02)* |
| | | *"As councilor I noticed the construction material was not enough, so we mobilized transport the National park we asked for a tractor to help us carry sand, we also asked Ebenezer they are close by here so they also helped in carrying the sand." (Councillor, Male, Clinic 01)* |
| | | *"It (Fambidzanai) helped with roofing sheets and food for builders, whilst World Vision Zimbabwe helped with windows, roofing sheets, window frames, cement and everything that is needed for construction…and those in the diaspora helped with paying the builders and buying food for them." (Community member, Female, Clinic 01)* |
| | | *"We then agreed as a society group that the monthly contributions we made toward burial plans we were going to channel them toward the development building project." (Key informant 06, Male, Clinic 03)* |
| Barriers to community participation | Lack of interest, distrust, and resistance to local leadership | "They were refusing, some said we have never seen a clinic being built from mud bricks, some would say I won't waste my time going for the construction and others said it was all politics so we don't do that, till the |

| | | |
|---|---|---|
| | | clinic got finished some didn't even come not even a single day…They are the ones' in the fore front now (laughs)." (Community member, Female, Clinic 01) |
| | | *"The challenges that we faced is that the people that would work will be a few and yet we have eight hundred and forty something in these wards… We couldn't get anyone to assist these people (builders) if they spent the day at work, they wouldn't get food…and at times we failed to raise money to pay them." (Key informant, Clinic 02, Male)* |
| | | *"When we started many people were in support of the idea but practically, they did not show up in numbers because when it comes to manual labour and things that need money people tend to not cooperate." (Key informant 06, Clinic 03, Male)* |
| | | *"What I can say is that what affects other regions is politics. People should focus more on development and not politics. Those in power, like chiefs, should also coordinate others they shouldn't be power greedy" (Key informant, Clinic 02, Male)* |
| | Financial constraints | *"people agreed to contribute as per household and each household was supposed to contribute 20 US dollars or the* |

| | | |
|---|---|---|
| | | *equivalent of that in bond Zimbabwean currency. Unfortunately, it was not a success because people had financial challenges." (Key informant_08, Clinic 02, Male)* |
| | | *"We had a challenge contributing money to give the builders. The council had promised that they will give them wages, but they failed to raise them and at the end of the day they stopped" (Key informant, Clinic 02, Male)* |
| | | *"everyone who is a builder got their names noted down…but when it came to action only a few came (both laugh) because of influences from others and if for instance, a wife complain that they are suffering from hunger at home whilst he spent time at the clinic working with no wage those men ended up retreating leaving only a few." (Community member, Clinic 01, Female)* |
| Positive outcomes of the VHSM | Improved health outcomes | *"People would get sick and some would not rush to the clinic because of the distance and expenses, hence they would end up getting more complications. Now the greatest advantage we have is that we have reduced number of illnesses and even complications because people get help faster. Now they are reduced home deliveries because back then some women will* |

| | | |
|---|---|---|
| | | *delay going to the hospital and end up giving birth along the way to hospital." (Key informant_08, Clinic 02, Male)* |
| | | *"As of now there are no defaulters anymore because a person would fail to get someone to send and get tablets for them at the clinic so without them they were forced to default, so people couldn't access tablets because of money for transport then spend the whole week without taking tablets, but now it's easier because the clinic is now near." (Community member, Clinic 01, Female).* |
| | | *"The number of deaths in children has greatly reduced, because children would die because they were not immunized." (Village Health Worker 04, Clinic 03, Female)* |
| | Sustainability of community led initiatives | *"This initiative has made people work together. It has motivated people and as of now they want to construct a cottage for the nurses because they have realised working together works. Thanks to that they have learned that working together is an easier way they can fend for themselves." (Key informant, Clinic 02, Male)* |

| | | |
|---|---|---|
| Recommendations for future development plans | | *"As a community you need to sit down and raise all the concerns you have because at the end of the day it's you who suffer by walking long distances for a service that you can build when you unite and gather resources." (Key informant_08, Clinic 02, Male)* |
| | Capacity building of locals | *"On record keeping there is a need for people to be educated in all committees so they know the importance of record keeping, because if we look deep into it when I arrived some materials had arrived already but to trace it back to who contributed and gave a hand for instance the diaspora but it could not be traced. So on transparency there is still a challenge there is need for trainings." (Councillor, Clinic 01, Male)* |
